# Severity Assessment and Progression Prediction of COVID-19 Patients based on the LesionEncoder Framework and Chest CT

**DOI:** 10.1101/2020.08.03.20167007

**Authors:** You-Zhen Feng, Sidong Liu, Zhong-Yuan Cheng, Juan C. Quiroz, Dana Rezazadegan, Ping-Kang Chen, Qi-Ting Lin, Long Qian, Xiao-Fang Liu, Shlomo Berkovsky, Enrico Coiera, Lei Song, Xiao-Ming Qiu, Xiang-Ran Cai

## Abstract

Automatic severity assessment and progression prediction can facilitate admission, triage, and referral of COVID-19 patients. This study aims to explore the potential use of lung lesion features in the management of COVID-19, based on the assumption that lesion features may carry important diagnostic and prognostic information for quantifying infection severity and forecasting disease progression.

A novel LesionEncoder framework is proposed to detect lesions in chest CT scans and to encode lesion features for automatic severity assessment and progression prediction. The LesionEncoder framework consists of a U-Net module for detecting lesions and extracting features from individual CT slices, and a recurrent neural network (RNN) module for learning the relationship between feature vectors and collectively classifying the sequence of feature vectors.

Chest CT scans of two cohorts of COVID-19 patients from two hospitals in China were used for training and testing the proposed framework. When applied to assessing severity, this framework outperformed baseline methods achieving a sensitivity of 0.818, specificity of 0.952, accuracy of 0.940, and AUC of 0.903. It also outperformed the other tested methods in disease progression prediction with a sensitivity of 0.667, specificity of 0.838, accuracy of 0.829, and AUC of 0.736. The LesionEncoder framework demonstrates a strong potential for clinical application in current COVID-19 management, particularly in automatic severity assessment of COVID-19 patients. This framework also has a potential for other lesion-focused medical image analyses.

## 1. Introduction

The rapid escalation in the number of COVID-19 infections exceeded the capacity of healthcare systems to respond in many nations, and consequently reduced patient outcomes [1]. In such circumstances, it is of paramount importance to develop efficient diagnostic and prognostic models for COVID-19, so that the patients’ care can be optimised.

Chest CT scans have been found to provide important diagnostic and prognostic information for COVID-19 [2-7]. Although there is still debate on the use of chest CT in screening and diagnosing COVID-19 cases [8], a surge of computational methods for chest CT have been developed to support medical decision making during the current pandemic [9-15]. Study population, model performance, and reporting quality vary substantially between studies. An in-depth comparison of these studies can be found in a recent systematic review [16].

In addition to diagnostic and screening models, several prediction models have been proposed based on an assessment of lung lesions. There are three typical classes of lesions that can be detected in COVID-19 chest CT scans: ground glass opacity (GGO), consolidation, and pleural effusion [3,4]. Imaging features of the lesions including shape, location, extent and distribution of involvement of each abnormality, have been found to have good predictive power for mortality [17] or hospital stay [18]. These features, however, are mostly derived from the delineated lesions, and so depend heavily on lesion segmentation. Manual delineation of lesions often takes 1 to 5 hours, which substantially undermines clinical applicability of these methods.

Automatic lung lesion segmentation for COVID-19 has been actively investigated in recent studies [19,20]. A VB-Net model based on a neural network was proposed to segment the infection regions in CT scans [19]. This model, when trained using CT scans of 249 COVID-19 patients, achieved a Dice score of 0.92 between automatic and manual segmentations, and successfully reduced the delineation time to less than 4 minutes. In another recent study [20], a lesion segmentation model based on the 3D-Dense U-Net architecture was proposed and trained on CT scans of a combination of 160 COVID-19, 172 viral pneumonia, and 296 interstitial lung disease patients. Although the lesion masks were not compared voxel-to-voxel, the volumetric measures of lesions, such as percentage of opacity and consolidation, showed a high correlation (0.97-0.98) between automatic and manual segmentations.

Previous studies [19,20] have suggested that lesion features might be a useful biomarker for COVID-19 patient severity assessment, but the effectiveness of lesion features is yet to be verified. Lesion features may have additional applications in the management of COVID-19, which need to be investigated further. In this study, we aim to test the effectiveness of using lesion features in COVID-19 patients for disease severity assessment, and to explore the potential use of lesion features in predicting disease progression.

Automatic severity assessment and progression prediction will substantially facilitate admission, triage, and referral of patients. The first goal of this study is to develop a method for *assessing severity* of COVID-19 patients based on their baseline chest CT scans. Four severity types: mild, ordinary, severe, and critical, can be defined based on a core outcome set (COS) encapsulating clinical symptoms, physical and chemical detection, viral nuclei aid detection, disease process, etc. [21]. Supportive treatments, such as supplementary oxygen and mechanical ventilation, are usually required for severe and critical cases [22]. We represent the assessment severity task as a binary classification problem, i.e., to classify a patient as a mild / ordinary case (mild class) or a severe / critical case (severe class).

The second goal of this study is to *predict disease progression* for the mild / ordinary cases based on their baseline CT scans. In other words, we aim to predict which of the mild/ordinary severity patients are likely to progress to the severe / critical category (converter class) in the short term (within 7 days), and which patients would remain stable or recover (non-converter class), based on the assumption that lesion features may carry important prognostic information for forecasting disease progression. We again consider the task as a binary classification problem, i.e., to classify the non-converter cases and converter cases. Figure 1(a) presents an example of a COVID-19 case with mild symptoms. In less than 7 days, the patient’s symptoms rapidly worsened and progressed to severe. Figure 1(b) is an example of a non-converter case whose symptoms progressed slowly and remained mild 7 days after the baseline CT scan.

**Figure 1.**
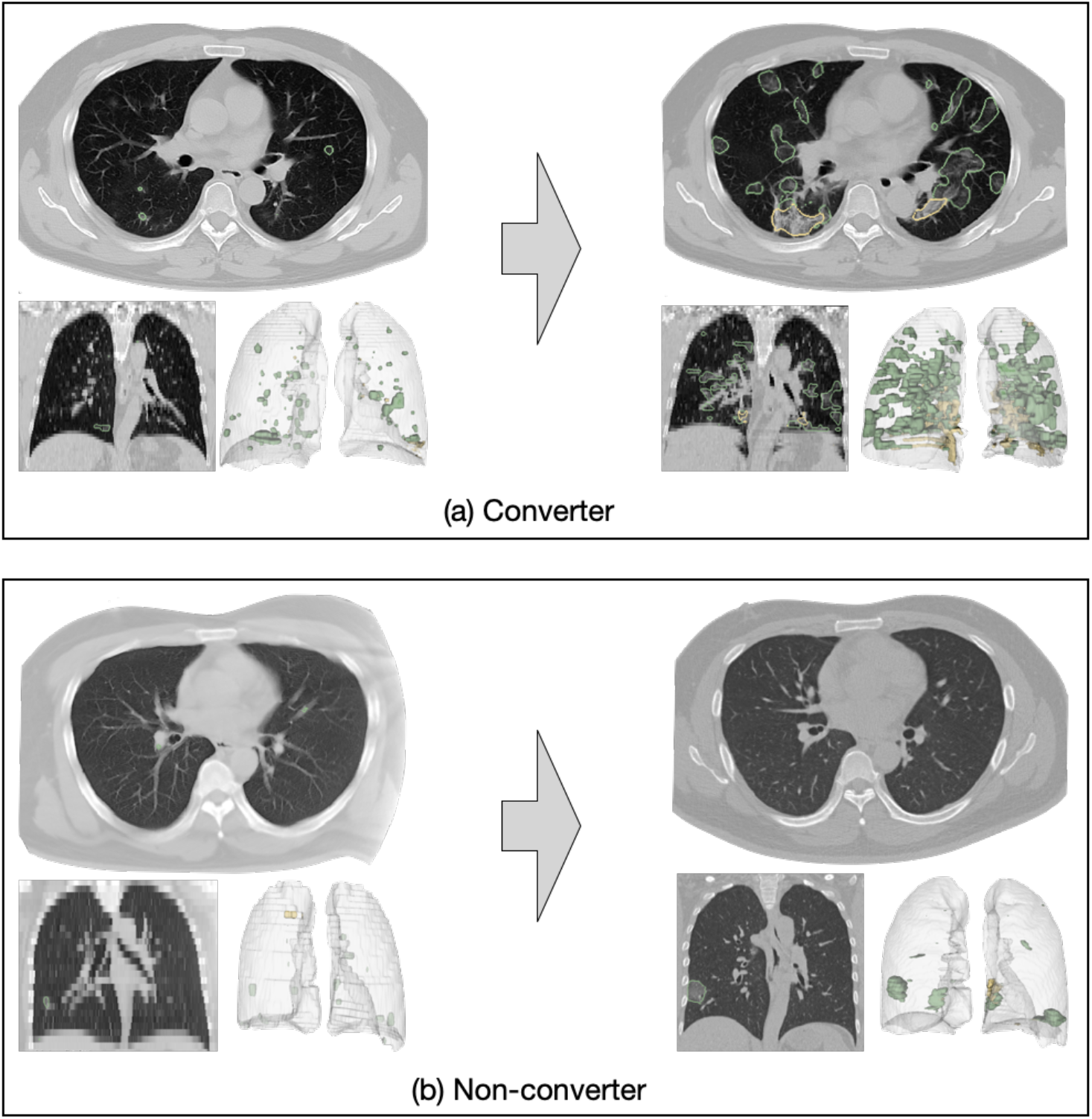
Examples of converter and non-converter cases. (1) a mild case progressed to severe within 7 days; (2) a mild case did not progress to severe within 7 days.

To achieve the above two goals, a novel **LesionEncoder** framework is proposed to detect lesions in CT scans and encode lesion features for automatic severity assessment and progression prediction. The **LesionEncoder** framework consists of two modules: (1) a U-Net module which detects lesions and extracts features from CT slices, and (2) a recurrent neural network (RNN) module for learning the relationship between feature vectors and classifying the sequence of feature vectors as a whole.

We applied the **LesionEncoder** framework for both severity assessment and progression prediction. With access to data of two COVID-19 confirmed patient cohorts from two hospitals, we trained our proposed model with CT scans of a cohort of patients from one hospital and tested it on an independent cohort from the other hospital. The models built on the **LesionEncoder** framework outperformed the baseline models that used lesion volumetric features and general imaging features, demonstrating a high potential for clinical applications in the current COVID-19 management, particularly in automatic severity assessment of COVID-19 patients. This framework may also have a strong potential in similar lesion-focused analyses, such as neuroimaging based brain tumor grading and retinal imaging based diabetic retinopathy grading.

## 2. Datasets

A total of 346 COVID patients confirmed by reverse transcription polymerase chain reaction (RT-PCR) were retrospectively selected from two local hospitals in the Hubei Province, China, namely Huang Shi Central Hospital (HSCH) and Xiang Yang Central Hospital (XYCH). Severity types of all patients at baseline and follow-up (in 7 days) were assessed and confirmed by clinicians according to the COS for COVID-19 [21]. This analysis was approved by the Institutional Review Board of both hospitals, and written informed consent was obtained from all the participants.

Tables 1 and 2 illustrate respectively the demographics of patients for the development of a severity assessment model (**Task 1 – mild vs severe**) and a progression prediction model (**Task 2 - converter vs non-converter**). For both tasks, CT scans of the HSCH cohort were used for training the models, and CT scans of the XYCH cohort were used as an independent dataset to test the trained models. Patients may have either a lung-window scan, a mediastinal-window scan, or both in their baseline CT examination. All scans were included in the analysis. The total number of CT scans for **Task 1** was 639, and that for **Task 2** was 601. An internal validation set (20% of the training samples) was split from the training set and used to evaluate the model’s performance during training.

**Table 1.** Demographics ^ of the patients in Task 1 dataset.

| Category | HSCH - Training Set | XYCH - Test Set | Total |
| --- | --- | --- | --- |
| Mild | 7 | 1 | 8 |
| Ordinary | 212 | 104 | 316 |
| Severe | 7 | 6 | 13 |
| Critical | 4 | 5 | 9 |
| Total patients | 230 | 116 | 346 |
| Total CT scans | 433 | 206 | 639 |
| Age (mean±SD) | 49.00±14.4 | 47.5±17.2 | 48.5±15.4 |
| Gender (female/male) | 120/110 | 57/59 | 177/169 |

**Table 2.** Demographics of the patients in Task 2 dataset.

| Category | HSCH- Training Set | XYCH- Test Set | Total |
| --- | --- | --- | --- |
| Non-Converter | 201 | 99 | 300 |
| Converter | 18 | 6 | 24 |
| Total patients | 219 | 105 | 324 |
| Total CT scans | 412 | 189 | 601 |
| Age (mean±SD) | 48.4±14.0 | 46.1±16.6 | 47.7±14.9 |
| Gender (female/male) | 113/106 | 55/50 | 168/156 |

Note that there is a highly imbalanced distribution of samples in the datasets, i.e., 324 (93.6%) patients in mild class for Task 1, and 300 (92.6%) patients in non-converter class for Task 2. A weighting strategy was used to address the imbalanced distribution in datasets, and the details are presented in Section 3.3.

## 3. Methods

Figure 2 gives an overview of the **LesionEncoder** framework, which consists of two modules: (1) a lesion encoder module for lesion detection and feature encoding, and (2) a RNN module for sequence classification. The lesion encoder module extracts features from individual CT slices; therefore, a CT scan with multiple CT slices can be represented as a sequence of feature vectors. The sequence classification module takes the sequence of feature vectors as input and then classifies the entire sequence collectively.

**Figure 2.**
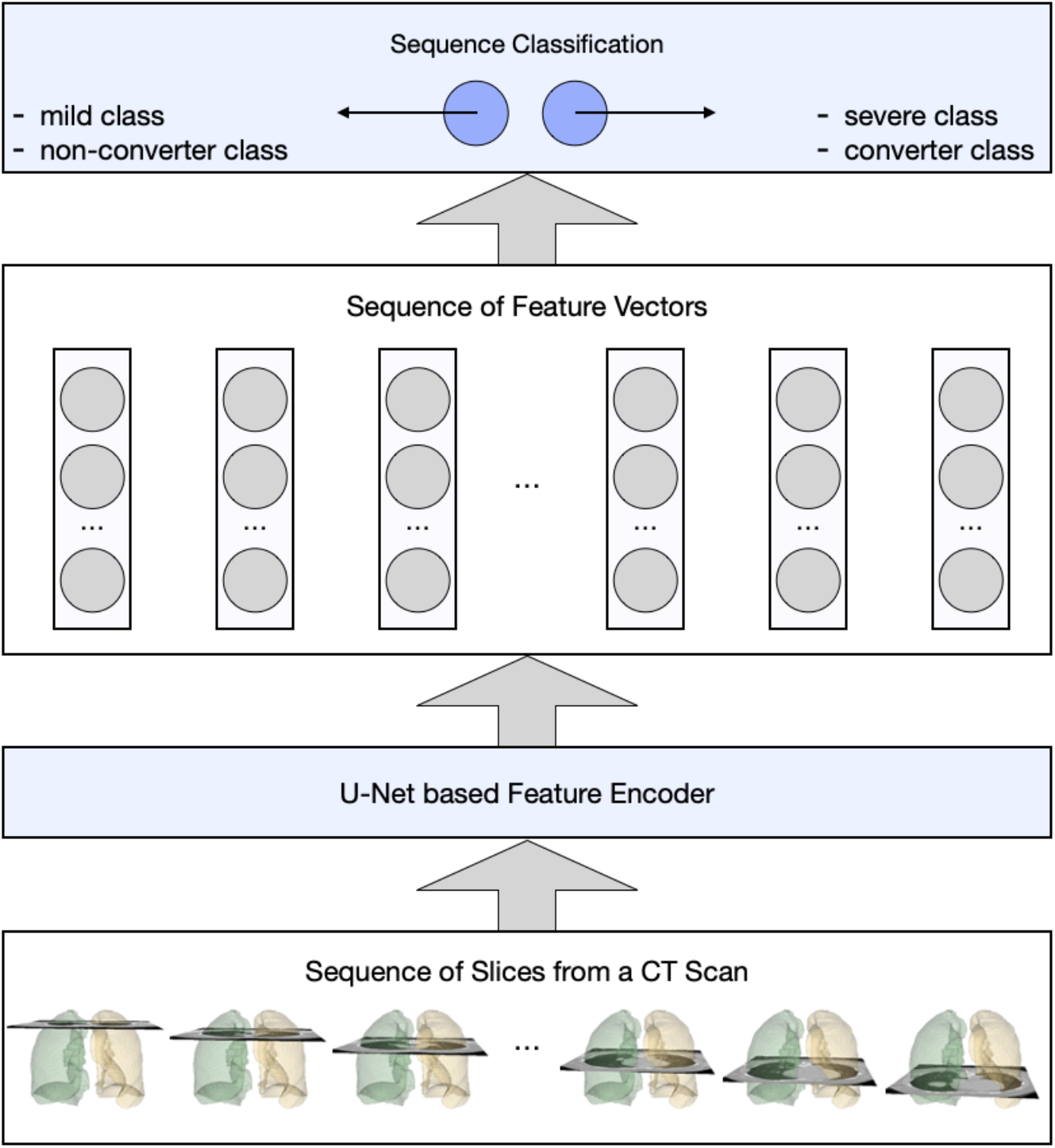
An overview of the proposed LesionEncoder framework.

### 3.1 Image Pre-processing

All CT scans were pre-processed with intensity normalization, contrast limited adaptive histogram equalization, and gamma adjustment, using the same pre-processing pipeline as in our previous study [23]. We further performed lung segmentation on the CT slices using an established model - R231CovidWeb [24]. This model^1^ was trained on a large and diverse dataset of non-COVID-19 CT scans and further fine-tuned with an additional COVID-19 dataset [25]. The CT slices with less than 3mm^2^ lung tissue were removed from our datasets, since they bear little or no information of the lung.

### 3.2 Lesion Encoder

The U-Net architecture [26] is adopted for the lesion encoder module. It consists of an encoder and a decoder, where the encoder captures the lesion features and the decoder maps lesion features back to the original image space. In other words, the encoder is responsible for extracting features from the input images, i.e., CT slices, whereas the decoder generates the segmentation maps, i.e., lesion masks. Figure 3 illustrates the encoder-decoder architecture of the lesion encoder module.

**Figure 3.**
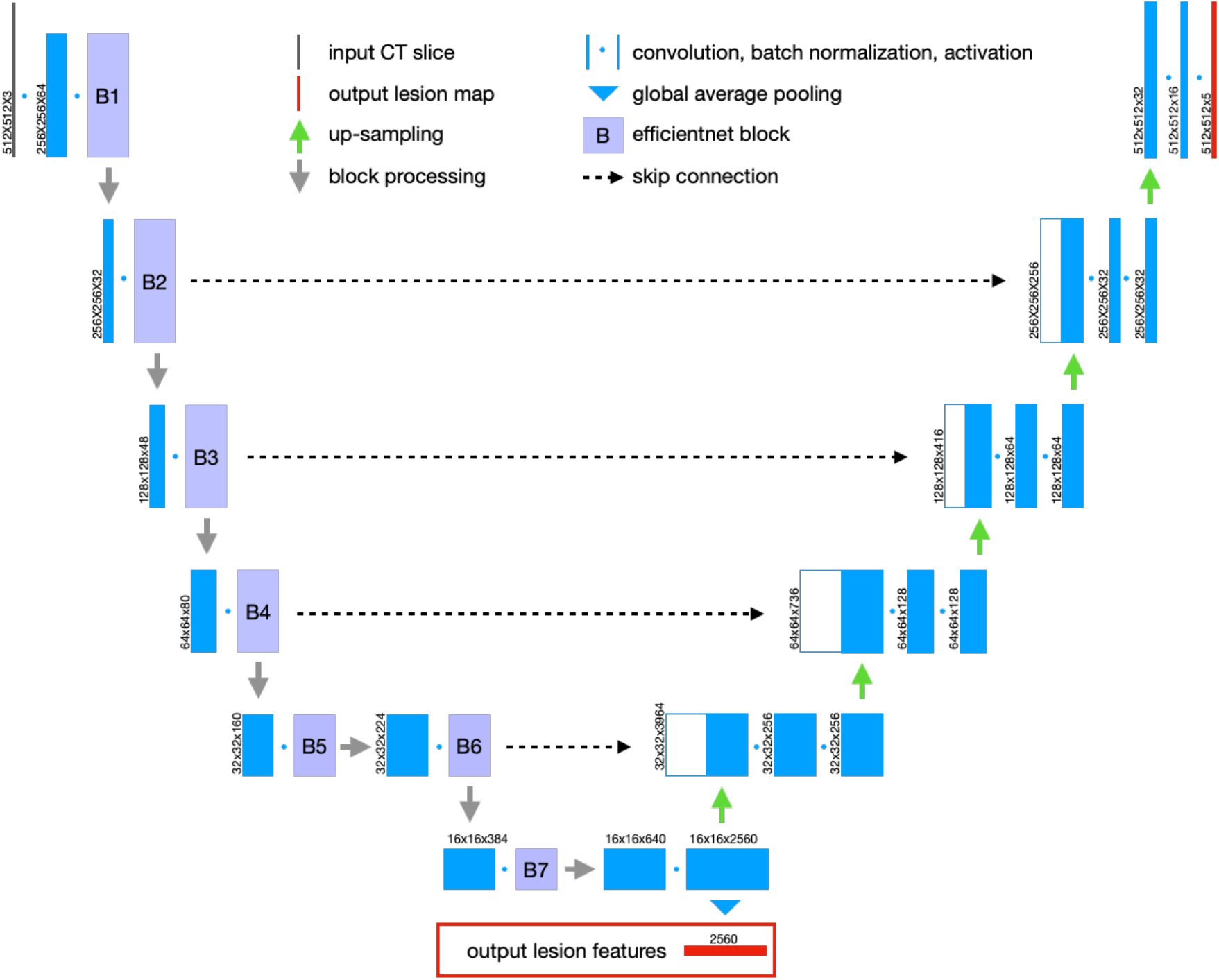
the U-Net architecture for lesion detection and feature encoding.

We used the EfficientNetB7 model [27] as the backbone to build the lesion encoder module, as it represents the state-of-the-art in object detection while being 8.4 times smaller and 6.1 times faster on inference than the best existing models in the ImageNet Challenge [28]. The ImageNet pre-trained weights were used to initialize the EfficientNetB7 model. There are 7 blocks in the EfficientNetB7 model, as shown in Figure 3. The skip connections were built between the expand activation layers in Block 2, 3, 4 and 6 and their corresponding up-sampling layers in our model. The output of the bottom layer is the final output feature vector representing the lesion features of the input slice.

A publicly available dataset was used to train the EfficientNetB7 U-Net, which consisted of 100 axial CT slices from 60 COVID-19 patients [25]. All the CT slices have been annotated by an experienced radiologist with 3 different lesion classes, including GGO, consolidation, and pleural effusion. Since this dataset is very small, we applied different augmentations, including horizontal flip, affine transforms, perspective transforms, contrast manipulation, image blurring and sharpening, Gaussian noise, and random crops, to the dataset using the Albumentations library [29]. The model^2^ was trained using Adam optimizer [30] with a learning rate of 0.0001 and 300 epochs.

The lesion encoder module was applied to process individual slices in a CT scan. For each CT slice, a high-dimensional feature vector (d=2,560) was derived. Independent component analysis (ICA) was performed on the training samples to reduce dimensionality (d=64). The ICA model was then applied to the test samples, so that they have the same feature dimension as the training samples. The output of the lesion encoder is a sequence of feature vectors, which are then classified using a sequence classifier, as explained in the next section.

### 3.3. Sequence Classification

A RNN model was built for sequence classification. Its input is a sequence of feature vectors generated by the lesion encoder. The structure of the RNN model is illustrated in Table 3 – two bidirectional Long Short-Term Memory (LSTM) layers, followed by a dense layer with a dropout rate of 0.5, and an output dense layer. For comparison purposes, another Pooling model was created (Table 3) – using max pooling and average pooling to combine the slice-based feature vectors, as inspired by a previous study [9]. The difference between these two models is that the RNN model captures the relationship between feature vectors in a sequence, whereas the Pooling model ignores such relationships.

**Table 3.** The architectures of the RNN model and the Pooling model.

| RNN Model | Pooling Model |  |
| --- | --- | --- |
| BiLSTM (64, return-sequences) | Global_Max_Pooling | Global_Average_Pooling |
| BiLSTM (32) | Concatenation |  |
| Dense (64, ReLu, dropout = 0.5) | Dense (64, ReLu, dropout = 0.5) |  |
| Dense (1, Sigmoid) | Dense (1, Sigmoid) |  |

Adam optimizer [30] with a learning rate of 0.001 was used for training the models in 100 epochs. A validation set (20%) was split from the training set for monitoring the training process. Every 20 epochs, the validation set was reselected from the training set, so that the model will be internally validated by all training samples during training. To address the imbalanced distribution in the datasets, we assigned different weights to the two classes (mild / non-converter class: 0.2, severe / converter class: 1.8) when training the models. In addition, if a patient has multiple CT scans, the scan with a higher probability of a positive prediction overrules the others when applying the models for inference.

### 3.4 Performance Evaluation

We tested the **LesionEncoder** framework with two configurations: (1) using the Pooling model as the classifier (**LE_Pooling**) and (2) using the RNN model as the classifier (**LE_RNN**).

These methods were compared to 3 baseline methods. The first baseline method (**BS_Volumetric**) was inspired by a previous study [20], which was based on a Logistic Regression model using 4 lesion volumetric features as input: GGO percentage, consolidation percentage, pleural effusion percentage, and total lesion percentage. The second (**BS_Pooling**) and third (**BS_RNN**) baseline methods were based on the same classification models as in LE_Pooling and LE_RNN; however, the features were extracted from an EfficientNetB7 model without a lesion encoder module. The purpose of the second and third baseline models was to estimate the contribution of the lesion encoder. Sensitivity, specificity, accuracy, and area under curve (AUC) were used to evaluate the methods’ performance. Receiver operating characteristic (ROC) curves were also compared between methods.

### 3.5 Development Environment

All the neural network models, including the EfficientNetB7 U-Net, the Pooling model and the RNN model, were implemented in Python (v3.6.9) and Tensorflow (v2.0.0). The models were trained using a Fujitsu server with Intel Xeon Gold 5218 GPU, 128G memory, and NVidia V100 32G GPU. The same server was used for image pre-processing, feature extraction, and classification.

## 4. Results

### 4.1 Lung and Lesion Segmentation

The lung masks generated using the R231CovidWeb model [24] and the lesion masks generated by the lesion encoder module were visually inspected by an experienced image analyst (S.L.). Overall, the lung segmentation results were visually reliable with few severe and critical cases having infection areas missed out in their lung masks. The lesion encoder achieved a Dice of 0.92 on the COVID-19 CT segmentation dataset [25]. Figure 4 presents 4 examples of the lung and lesion segmentation results of the COVID-19 patients, one for each severity class. The upper row presents the axial CT slices with the lung (red) and lesion (green: GGO; yellow: consolidation; brown: pleural effusion) boundaries overlaid on the CT slices.

**Figure 4.**
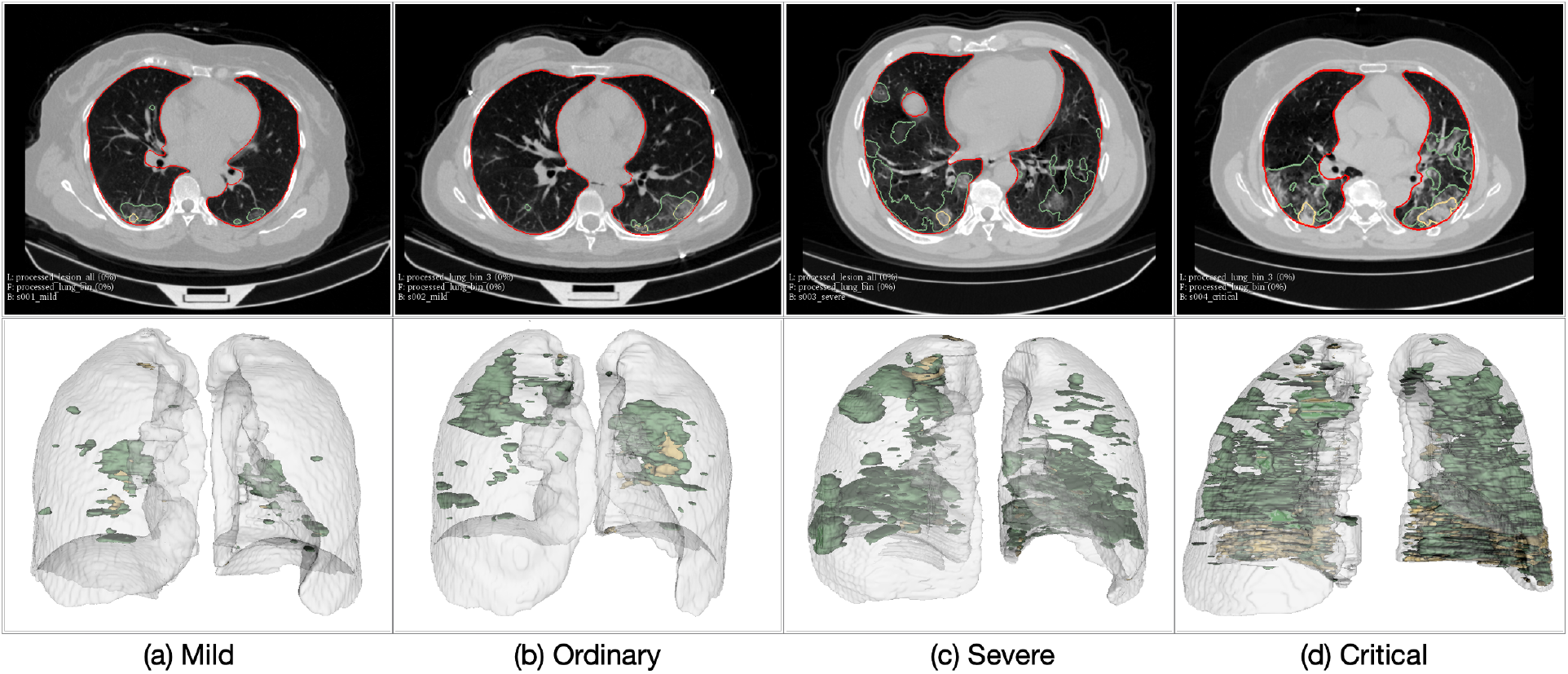
Examples of the patients in different severity groups.

The lower row of Figure 4 illustrates the 3D models of the lung and lesions, reconstructed using 3D Slicer (v4.6.2) [31]. Figure 4 shows that higher severity of COVID-19 is reflected in CT scans as increasing number and volume of lesions.

### 4.2 Severity Assessment

Five different methods were compared in the automatic severity assessment of COVID-19 patients, including 3 baseline methods and 2 proposed methods, as described in Section 3.4. Table 4 illustrates the performance metrics of different methods on the severity assessment task, and Figure 5(a) shows the ROC curves of these methods. The three methods using lesion features (BS_Volumetric, LE_Pooling, and LE_RNN) consistently outperformed the models that did not use lesion features by a marked difference in sensitivity (>9.1%), specificity (>15.3%), accuracy (>14.7%), and AUC (>15.1%). In particular, BS_Volumetric achieved the highest AUC of 0.931, indicating that the lesion volumetric features were highly effective in distinguishing between severe and mild cases.

The proposed LE_RNN method achieved higher specificity (0.952) than the BS_Volumetric method (0.933), showing that the features captured by the lesion encoder might be useful in reducing the false positive rate compared with the volumetric features. When comparing the Pooling models and RNN models, we found that the RNN models performed slightly better than the Pooling models; and the impact of the sequence classifier on the classification performance was much lower than that of the lesion features.

**Table 4.** Performance of different methods in baseline severity assessment.

| Method | Sensitivity | Specificity | Accuracy | AUC |
| --- | --- | --- | --- | --- |
| BS_Volumetric | **0.818** | 0.933 | 0.922 | **0.931** |
| BS_Pooling | 0.727 | 0.752 | 0.750 | 0.732 |
| BS_RNN | 0.727 | 0.771 | 0.767 | 0.749 |
| **LE_Pooling** | **0.818** | 0.924 | 0.914 | 0.900 |
| **LE_RNN** | **0.818** | **0.952** | **0.940** | 0.903 |

### 4.3 Progression Prediction

The results of different methods in the prediction of disease progression task are presented in Table 5 and Figure 5(b) presents the ROC curves of these methods. The BS_Volumetric method performed poorly (sensitivity: 0.5, specificity: 0.465, accuracy: 0.467, AUC: 0.51), indicating that lesion volumetric features were not predictive of COVID-19 disease progression. This finding was not surprising, since the converter and non-converter cases both showed mild symptoms at baseline and presented a small quantity of lesions in the lungs. The BS_Pooling and BS_RNN methods achieved slightly better performance than BS_Volumetric, although they did not use any lesion features.

The LE_Pooling and LE_RNN methods outperformed the baseline methods with a substantial increase of 20-30% in specificity. LE_RNN was the best method in all the evaluation metrics (sensitivity: 0.667, specificity: 0.838, accuracy: 0.829, AUC: 0.736). The results indicate that the lesion features extracted by the lesion encoder may bear useful diagnostic information for predicting disease progression. However, it is still challenging to predict disease progression using the lesion features, and the low sensitivity (0.667) may restrict clinical applicability of the proposed methods.

**Table 5.** Performance of different methods in prediction of disease progression.

| Method | Sensitivity | Specificity | Accuracy | AUC |
| --- | --- | --- | --- | --- |
| BS_Volumetric | 0.500 | 0.465 | 0.467 | 0.510 |
| BS_Pooling | **0.667** | 0.535 | 0.543 | 0.569 |
| BS_RNN | **0.667** | 0.535 | 0.543 | 0.662 |
| **LE_Pooling** | **0.667** | 0.737 | 0.733 | 0.724 |
| **LE_RNN** | **0.667** | **0.838** | **0.829** | **0.736** |

**Figure 5.**
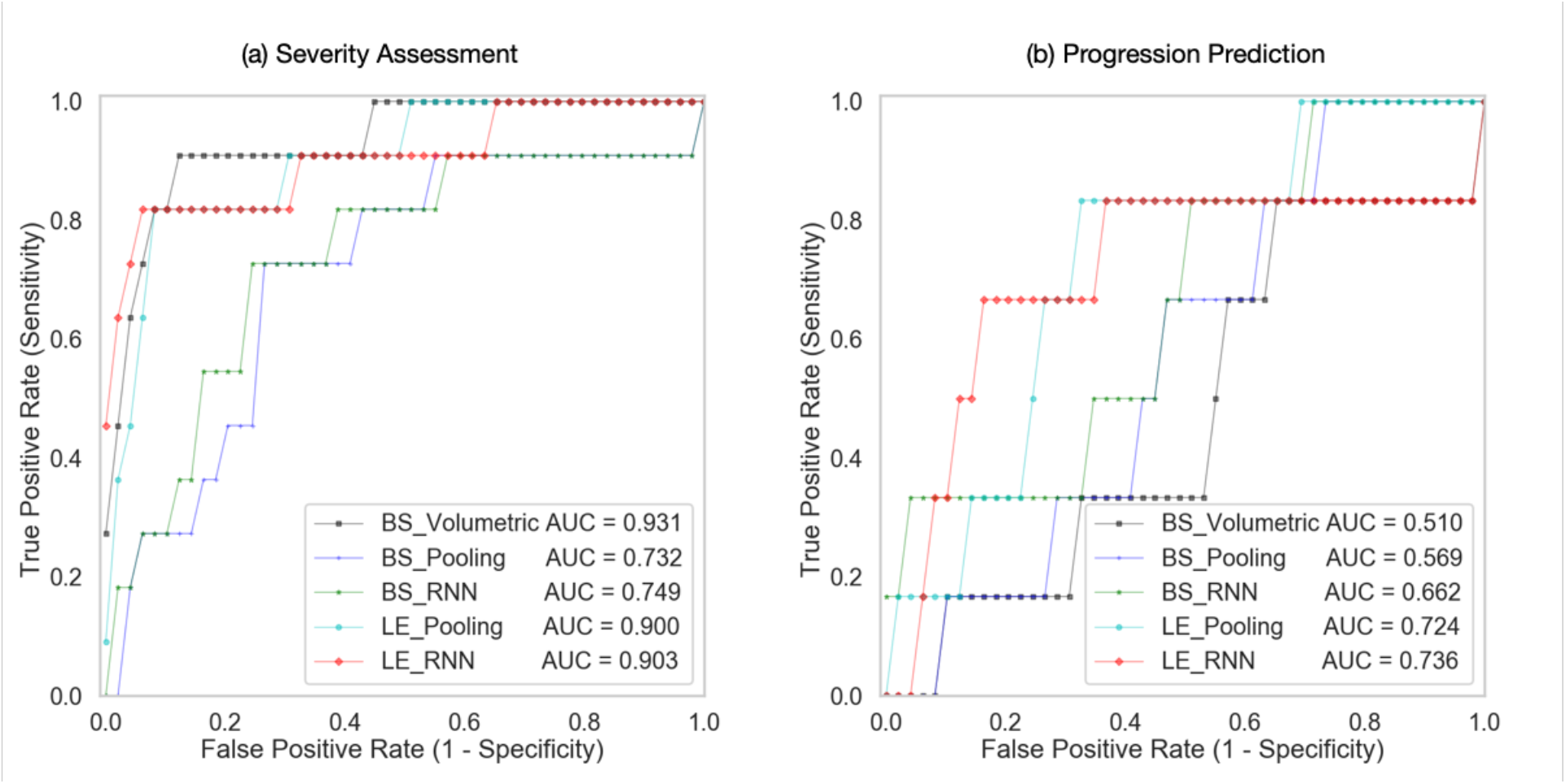
ROC curves of different models in severity assessment and progression prediction.

## 5. Discussion

### Clinical value in the management of COVID-19

The rapid spread of COVID-19 has put a strain on healthcare systems, necessitating efficient and automatic diagnosis and prognosis to facilitate the admission, triage, and referral of COVID-19 patients. Chest CT plays a key role in COVID-19 management by providing important diagnostic and prognostic information of patients. Several computational models have been developed to support automatic screening and diagnosis of COVID-19 [9-15]. There are also a few studies [19,20] using CT to quantify infection severity with a focus on development of lesion segmentation models. A few measures based on the lesion volumes have been proposed to quantify infection severity [19], however, the intricate patterns of the lesion shape, texture, location, extent and distribution, were less investigated.

To capture the complex features in the lesions, we proposed a novel **LesionEncoder** framework. Two specific applications of this framework, i.e., assessment of severity and prediction of disease progression for COVID-19 patients, were explored in this study. To the best of our knowledge, this work represents the first attempt to predict COVID-19 patient disease progression using chest CT scans. Models built on this framework are able to take CT scans as input, detect and extract features from the lesions, and quantify the severity or predict progression in a fully automated manner. The analysis of a high-resolution CT scan of 512×512×430 voxels takes less than 1 minute, which is substantially faster than radiologists’ reading time. This can also save the burden of manual delineation of the lesions. The quantitative measures based on the features are of high clinical relevance, and can be used to support medical decision making or to track changes in patients.

We should note that this framework is not designed to analyse the COVID-19 suspects who are not confirmed by RT-PCR, or the covert / asymptomatic cases that are not documented [32-34]. The community-acquired pneumonia cases, such as viral pneumonia and interstitial lung disease patients, were also not considered in this study. As pointed out in a systematic review [35], normal controls and diseased controls will be needed for the development of screening or diagnostic models, thus the selection bias in cohort may lead to a risk of overestimated performance. Since our model focuses on the confirmed and hospitalized COVID-19 cases, therefore will not be exposed to such risk.

Models based on lesion features outperformed the baseline models without lesion features in both severity assessment and progression prediction. An interesting finding in severity assessment is that the lesion volumetric features are highly effective in distinguishing the severe cases from the mild cases. The features extracted by the lesion encoder did not improve the sensitivity of detecting the severe cases, but only reduced false positive predictions. This finding indicates that lesion volumetric features, such as GGO percentage and consolidation percentage, are prominent biomarkers in identifying the severe cases. The lesion encoder makes marginal contribution to severity assessment. In contrast, in progression prediction the models with the lesion encoder performed much better than those with volumetric features, indicating that the intricate pattern captured by the lesion encoder provide useful prognostic information for identifying the COVID-19 patients at higher risk of converting to the severe type. The **LesionEncoder** framework demonstrates a clinical applicability in COVID-19 management, particularly in the automatic severity assessment of COVID-19 patients (sensitivity: 0.818, specificity: 0.952, AUC: 0.931). However, it is still challenging to predict disease progression using lesion features, and the low sensitivity (0.667) may restrict clinical applicability of the proposed methods.

### Technical contributions of the LesionEncoder framework

The technical contributions of this work are two-fold. Most importantly, this framework extends the use of lesion features beyond conventional lesion segmentation and volumetric analysis. There is a wealth of information in the lesions including shape, texture, location, extent and distribution of involvement of the abnormality, that can be extracted by the lesion encoder. We demonstrated two novel applications of the lesion features in severity assessment and progression prediction. However, they also have a strong potential in a wide range of other clinical and research applications, such as supporting clinical decision making and providing insights of the pathological mechanism.

In addition, the proposed **LesionEncoder** framework attempts to address a common challenge in medical image analysis: how to reconcile local information and global information to improve medical image perception [36]. In this study, the slices from a CT scan were used as input for classification, but not every slice in the scan carries the same diagnostic / prognostic information. That is, the ground truth label of the entire scan cannot be propagated to label individual slices. For example, a CT slice with no lesion from a severe case might appear more ‘normal’ compared to a slice with some lesions from a mild case. Our proposed framework is a feasible approach to infer the holistic prediction with a focus on the analysis of region of interest. The RNN module in the framework is also a more sophisticated approach than the conventional feature fusion methods that use average pooling or max pooling to combine the local features. There are many analyses of the same nature, e.g., neuroradiologists may use features such as tumoral infiltration of surrounding tissues in MRI for tumor grading [37]; ophthalmologists may focus on lesions, such as haemorrhages and microaneyrysms, hard exudates and cotton-wool spots, when grading diabetic retinopathy [38]; and pathologists are more likely fixate on regions of highest diagnostic relevance when interpreting the biopsy whole slide images (WSI) [39]. The **LesionEncoder** framework may be generalizable to these lesion-focused medical image analyses.

### Limitations

Although we have access to 639 CT scans of 346 patients, it is still a relatively small dataset compared to other datasets for development of deep learning models. It also refuted the idea of developing 3D deep learning models for scan-based classification, since 3D models are usually more complicated than 2D models and have substantially more parameters, the small sample size will lead to undertrained models. In addition, there is a highly imbalanced distribution in the datasets. Among the 346 samples for the development of the severity assessment model, 324 (93.6%) patients were in the mild class. For the disease progression model, there are 300 (92.6%) patients in the non-converter class. Although this reflects the real distribution, it will be ideal to have more severe / converter class samples for training. To address this imbalance distribution problem, we used a class weighting strategy to give the positive class higher weight during training, and used a prediction weighting strategy during inference to enhance the prediction of the positive class if that patient has multiple scans. A larger sample size with more severe and converter cases in the datasets would help train more accurate and robust models as well as produce reliable performance estimates.

The lung masks generated using the R231CovidWeb model [24] and the lesion masks generated by the lesion encoder module were visually inspected by an experienced image analyst. The segmentation results were visually reliable, but the missed-out lung or lesion regions in the segmentation masks were noted in a few severe and critical cases. Since there were no lesion masks for our datasets, no quantitative analyses were performed to evaluate the automatic segmentation results. Further improvements can be made if the ground truth annotation of the lung and lesion can be provided to optimize the performance of the current lung segmentation model and lesion encoder module on our datasets.

## 6. Conclusions

In this study, a novel **LesionEncoder** framework was proposed to encode the enriched lesion features in chest CT scans for automatic severity assessment and progression prediction of COVID-19 patients. Models built on this framework outperformed the evaluated baseline models with a marked improvement. The lesion volumetric features were prominent biomarkers in identifying severe / critical cases, but intricate features captured by the lesion encoder were found effective in identifying the COVID-19 patients who have higher risks of converting to the severe or critical type. Overall, the **LesionEncoder** framework demonstrates a high clinical applicability in the current COVID-19 management, particularly in automatic severity assessment of COVID-19 patients.

An important future direction of this framework lies in the combination of clinical data and imaging data for better prediction performance, especially for the progression prediction, since clinical data may provide essential indicators of the clinical risks of the patients. Furthermore, the applications of the **LesionEncoder** framework to other types of lesion-focused analyses will be further investigated.

## Data Availability

The dataset is not publicly available due to patient privacy restrictions, but may be available from the corresponding author on reasonable request.

## Author Contributions

The project was initially conceptualized and supervised by X.R.C., X.M.Q. and L.S. The patient data and imaging data were acquired by Y.Z.F., Z.Y.C. and D.R. The analysis methods were designed and implemented by S.L. and J.C.Q. The data were analyzed by S.L. and J.C.Q. The research findings were interpreted by P.K.C., Q.T.L, L.Q., X.F.L, S.B. and E.C. All authors were involved in the design of the work. The manuscript was drafted by Y.Z.F., S.L and L.Q., and all authors have substantively revised it. All authors have reviewed and approved the submitted version.

## Acknowledgements

This project was supported by Natural Science Foundation of Guangdong Province (grant no.2017A030313901); Guangzhou Science, Technology and Innovation Commission (grant no.201804010239) and Foundation for Young Talents in Higher Education of Guangdong Province (grant no.2019KQNCX005). Dr Sidong Liu acknowledges the support of an Australian National Health and Medical Research Council grant (NHMRC Early Career Fellowship 1160760). We acknowledge Fujitsu Australia Limited for providing the computational resources for this study. Special thanks to Dr Eva Huang for proofreading this paper.

**Abbreviations**
RNN: - recurrent neural network
GGO: - ground glass opacity
COS: - core outcome set
LSTM: - long short term memory
RT-PCR: - reverse transcription polymerase chain reaction

## Footnotes

1 The binary executable software for the lung segmentation model is available online (https://github.com/JoHof/lungmask).

2 The Tensorflow implementation of the EfficientNetB7 U-Net is available online (https://github.com/qubvel/segmentation_models).

